# Clinical Benefits and Budget Impact of Lenzilumab plus Standard of Care Compared with Standard of Care Alone for the Treatment of Hospitalized Patients with COVID-19 in the United States from the Hospital Perspective

**DOI:** 10.1101/2021.10.06.21264651

**Authors:** Adrian Kilcoyne, Edward Jordan, Allen Zhou, Kimberly Thomas, Alicia N. Pepper, Dale Chappell, Miyuru Amarapala, Avery Hughes, Melissa Thompson

## Abstract

**Aims:** The study estimated the clinical benefits and budget impact of lenzilumab plus standard of care (SOC) compared with SOC alone in the treatment of hospitalized COVID-19 patients from the United States hospital perspective.

**Materials and Methods:** An economic model was developed to estimate the clinical benefits and costs for an average newly hospitalized COVID-19 patient, with a 28-day time horizon for the index hospitalization. Clinical outcomes from the LIVE-AIR trial included failure to achieve survival without ventilation (SWOV), mortality, time to recovery, intensive care unit (ICU) admission, and invasive mechanical ventilation (IMV) use. Base case costs included drug acquisition and administration for lenzilumab and hospital resource costs based on the level of care required. The inclusion of 1-year rehospitalization costs was examined in a scenario analysis.

**Results:** In the base case and all scenarios, treatment with lenzilumab plus SOC improved all specified clinical outcomes over SOC alone. Adding lenzilumab to SOC was also estimated to result in cost savings of $3,190 per patient in a population aged <85 years with CRP <150 mg/L and receiving remdesivir (base case). Per-patient cost savings were also estimated in the following scenarios: 1) aged <85 years with CRP <150 mg/L, with or without remdesivir ($1,858); 2) Black and African American patients with CRP <150 mg/L ($13,154); and 3) Black and African American patients from the full population ($2,763). In the full mITT population, a budget impact of $4,952 was estimated. When adding rehospitalization costs to the index hospitalization, a total per-patient cost savings of $5,154 was estimated.

**Conclusions:** The results highlight the clinical benefits for SWOV, ventilator use, time to recovery, mortality, time in ICU, and time on IMV, in addition to a favorable budget impact from the United States hospital perspective associated with adding lenzilumab to SOC for patients with COVID-19 pneumonia.

## Introduction

The coronavirus disease 2019 (COVID-19) pandemic has had a substantial cumulative societal and economic impact in the United States (US) and worldwide [1-3]. As of July 5, 2021, over 33 million cases and 603,000 deaths were reported in the US [4]. Data from the US Centers for Disease Control and Prevention (CDC) indicate that 8% of all individuals who developed COVID-19 between August 2020 and March 2021 required hospitalization [5]. Based on data from a large subset of US hospitals, 44.6% of patients hospitalized with COVID-19 required intensive care unit (ICU) admission and 12.5% required invasive mechanical ventilation (IMV) over the same time period [6].

The overall healthcare cost burden associated with the treatment of COVID-19 is substantial [7-9]. From the hospital perspective, providing care to patients with COVID-19 is associated with average per-patient costs ranging from a low of $14,325 USD for patients who do not require ICU admission to a high of $78,245 USD for patients who require both ICU admission and IMV [8]. Given the detrimental health outcomes and high economic burden among hospitalized patients with COVID-19, particularly those who require IMV, there remains a significant unmet need for safe, effective, and cost-effective treatment options [10-12].

Many adverse clinical outcomes among patients with COVID-19 are the result of an immunopathological process called cytokine storm [13,14]. This process is, in a significant part, mediated by granulocyte-macrophage colony-stimulating factor (GM-CSF), which activates and mobilizes myeloid cells, leading to dysregulated production of inflammatory cytokines and elevation of inflammatory markers (including C-reactive protein [CRP]) [13,15]. In patients with COVID-19, levels of GM-CSF–secreting T-cells and the extent of inflammatory cytokine production are directly correlated with lung tissue injury, disease severity, and ICU admission [14,16-18]. Therefore, GM-CSF has been identified as an important therapeutic target in COVID-19 [18].

Lenzilumab is a novel Humaneered^®^ anti-human GM-CSF monoclonal antibody that directly binds GM-CSF, thereby preventing its downstream signaling [13]. The Phase 3 LIVE-AIR trial (NCT04351152) is a randomized, double-blind, placebo-controlled study designed to evaluate early intervention with lenzilumab compared with placebo, both in combination with standard of care (SOC), in newly hospitalized patients with COVID-19 who have an oxygen saturation (SpO_2_) ≤94% on room air or require supplemental oxygen but have not progressed to IMV [13]. Preliminary results of the LIVE-AIR trial showed that lenzilumab significantly improved the likelihood of achieving survival without ventilation (SWOV)^1^ (sometimes referred to as ventilator-free survival) by day 28 compared with placebo [13]. Lenzilumab plus SOC was most efficacious as measured by SWOV in the groups of patients aged <85 years with CRP levels <150 mg/L, and patients aged <85 years with CRP levels <150 mg/L receiving remdesivir, compared with SOC alone [13,19]. C-reactive protein is an important marker of systemic inflammation and elevated levels of CRP are associated with poor clinical outcomes among patients with severe COVID-19 [13,20,21]; therefore, preliminary findings from the LIVE-AIR trial suggest that lenzilumab may be particularly effective when used as an early intervention in hospitalized patients.

Results from a post hoc analysis of the LIVE-AIR trial also suggest that Black and African American patients may exhibit the greatest response to lenzilumab, with a nearly 9-fold increase in SWOV among patients with CRP levels <150 mg/L [22]. This is notable as Black and African American persons have a 3-fold greater risk of hospitalization and 2-fold greater risk of death from COVID-19 [23]. The hyper-vulnerability in this population may be attributed to the low vaccination rates, the high prevalence of chronic illness (e.g., diabetes, hypertension, obesity), and the social determinants of health (e.g., socioeconomic status, healthcare access), all of which result in higher risk of infection, hospitalization, and death [23-26]. Despite the disproportional incidence of COVID-19 in Black and African American persons, racial minority groups are typically underrepresented in COVID-19 clinical trials [27]. However, Black and African Americans were well represented in LIVE-AIR, representing 14.8% of the trial population, which closely aligns with real-world demographics from the latest US Census (13.4%) [22,28].

Although clinical evidence indicates that lenzilumab is effective at improving SWOV in hospitalized patients with COVID-19 [13], its overall clinical and economic value from the hospital decision maker perspective has not been previously characterized. The purpose of this study was to estimate the clinical benefits and budget impact of lenzilumab plus SOC compared with SOC alone in the treatment of hospitalized COVID-19 patients from the US hospital perspective.

## Methods

### Model Structure and Calculations

An economic model was developed in Microsoft^®^ Excel to estimate the clinical benefits and budget impact of adding lenzilumab to SOC versus SOC alone for newly hospitalized patients with COVID-19 pneumonia. The model was developed for a US hospital perspective, with a 28-day time horizon for the index COVID-19 hospitalization (base case) and a 1-year time horizon for long-term costs (scenario). A 28-day time horizon for the index hospitalization was selected to align with data censoring in the LIVE-AIR trial (28 days following enrollment) [13]. A patient-level analysis was conducted to estimate the clinical benefits and costs for an average newly hospitalized patient with COVID-19 pneumonia. The model structure is presented in **Figure 1**.

**Figure 1.**
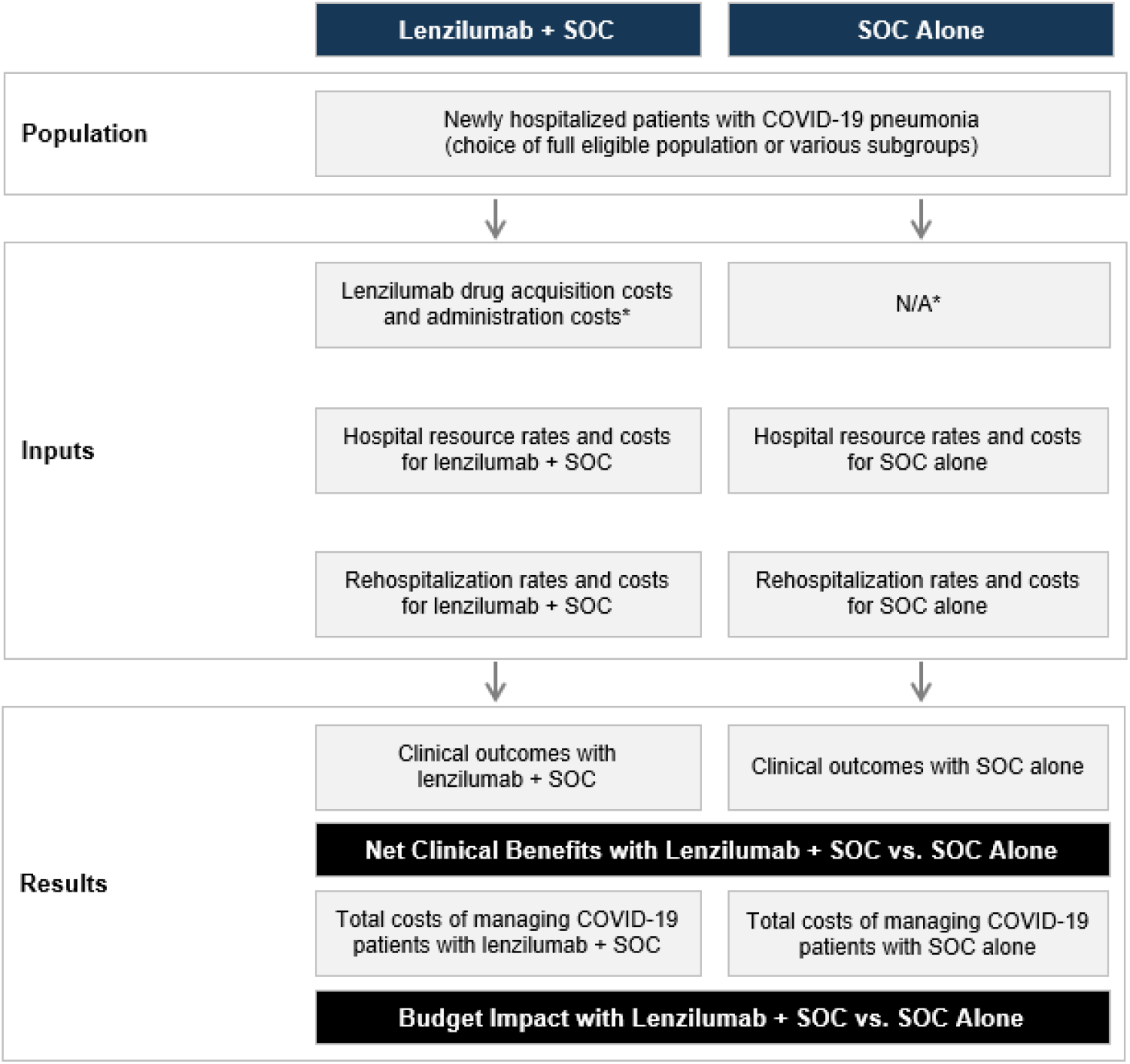
Model structure. Abbreviations. N/A, not applicable; SOC, standard of care. * It was assumed that use of SOC drugs would not be affected by the concomitant use of lenzilumab, resulting in no cost differences for SOC drugs between both groups. This assumption was supported by the balanced use of remdesivir and corticosteroids in both treatment arms of the LIVE-AIR trial [13]. ** Rehospitalization costs are optional in the model. They were not included in the base case analysis but were included for scenario analysis #5.

Clinical outcomes considered in the model included failure to achieve SWOV, mortality, time to recovery, ICU admission, and IMV use. Time to recovery^2^ was used as a proxy for a patient’s length of stay during the index hospitalization. Costs considered in the base case model included drug acquisition and administration costs for lenzilumab and hospital resource costs based on level of care required during the index hospitalization. The model also included the flexibility to explore the addition of rehospitalization costs within one year of the index hospitalization (scenario analysis). Adverse event (AE) management was not considered because the incidence of grade ≥3 AEs was similar in both arms of the LIVE-AIR trial (all differences for individual AEs between arms were less than 3%), and the lenzilumab arm had a lower overall incidence of grade ≥3 AEs (26.7% versus 32.7% for the safety populations of the lenzilumab and placebo arms, respectively) [13].

Based on a study by Di Fusco and colleagues [8] that examined the economic burden of hospitalized patients with COVID-19 in the US, patients were divided into four levels of care required: no ICU and no IMV, ICU but no IMV, IMV but no ICU, and both ICU and IMV. Hospital resource costs per patient were calculated as a weighted average based on the four levels of care required. First, the average time to recovery for each level of care was multiplied by the corresponding daily hospital resource use cost to obtain the average total hospital resource use cost for each level of care. Next, the weighted average cost per patient was calculated as the sum product of average total hospital resource use costs for each level of care and the corresponding proportion of patients requiring each level of care. These calculations were performed independently for the lenzilumab plus SOC and SOC alone arms to obtain separate weighted average costs for the two treatment options.

For long-term costs (scenario), only patients who survived the index hospitalization were eligible for rehospitalization. For IMV patients, this was calculated as the proportion with failure to achieve SWOV minus mortality after 28 days. For non-IMV patients, this was calculated as 100% minus the proportion with failure to achieve SWOV after 28 days.

### Model Inputs

#### Target Population

Patient eligibility for the model was based on inclusion criteria for the LIVE-AIR trial. In brief, eligible patients were newly hospitalized with COVID-19 pneumonia, with SpO_2_ ≤94% on room air and/or requiring supplemental oxygen, but not on IMV [13]. Data from the LIVE-AIR trial indicated that patients aged <85 years with CRP <150 mg/L particularly benefited from lenzilumab, and patients who received concomitant remdesivir also experienced greater benefit than patients who did not [13]. Furthermore, US clinical practice guidelines recommend remdesivir for treatment of hospitalized patients with COVID-19 who require supplemental oxygen but not IMV [29]. Therefore, a population that only included patients aged <85 years with CRP <150 mg/L and who were receiving remdesivir was deemed the most informative for hospital decision makers and was selected for the base case. This base case population is also in line with the population being evaluated in the ongoing ACTIV-5/BET-B trial that aims to further elucidate the efficacy of lenzilumab in this patient population [30]. Scenario analyses were also conducted in patients aged <85 years with CRP <150 mg/L (with or without remdesivir) (scenario #1), in the full LIVE-AIR modified intent-to-treat (mITT) population^3^ (scenario #2), in Black and African American patients with CRP <150 mg/L (with or without remdesivir) (scenario #3), and in Black and African American patients from the full mITT population (scenario #4). Data for the mITT population, the pre-specified primary analysis population, from the LIVE-AIR trial were used for all analyses [19].

#### Treatment Efficacy

Model inputs for treatment efficacy of lenzilumab plus SOC versus SOC alone were obtained from the LIVE-AIR trial [19]. Treatment efficacy inputs for the base case and scenario analyses are presented in **Table 1**.

**Table 1.** Treatment efficacy model inputs from LIVE-AIR trial data [19]^a^.

|  | Failure to achieve SWOV<sup>b</sup> | Mortality<sup>b</sup> | Time in ICU (days) | Time on IMV (days) |
| --- | --- | --- | --- | --- |
| **Base case: aged <85 years with CRP <150 mg/L, receiving remdesivir** |  |  |  |  |
| Lenzilumab plus SOC (*n* = 123) | 9.2% | 7.5% | 3.79 | 2.12 |
| SOC alone (*n* = 131) | 24.8% | 17.3% | 6.76 | 6.02 |
| **Scenario #1: aged <85 years with CRP <150 mg/L** |  |  |  |  |
| Lenzilumab plus SOC (*n* = 159) | 8.4% | 6.5% | 3.48 | 1.92 |
| SOC alone (*n* = 178) | 21.2% | 13.9% | 6.21 | 5.25 |
| **Scenario #2: full LIVE-AIR mITT population** |  |  |  |  |
| Lenzilumab plus SOC (*n* = 236) | 15.6% | 9.6% | 5.40 | 3.53 |
| SOC alone (*n* = 243) | 22.1% | 13.9% | 6.61 | 5.38 |
| **Scenario #3: Black and African American with CRP <150 mg/L** |  |  |  |  |
| Lenzilumab plus SOC (*n* = 25) | 4.0% | 4.0% | 1.48 | 1.12 |
| SOC alone (*n* = 26) | 29.3% | 17.1% | 6.54 | 6.62 |
| **Scenario #4: Black and African American** |  |  |  |  |
| Lenzilumab plus SOC (*n* = 38) | 13.2% | 7.9% | 4.68 | 3.11 |
| SOC alone (*n* = 33) | 29.1% | 16.5% | 7.06 | 6.48 |

Abbreviations. CRP, C-reactive protein; ICU, intensive care unit; IMV, invasive mechanical ventilation; mITT, modified intent-to-treat; SOC, standard of care; SWOV, survival without ventilation.
<sup>a</sup> All data were censored at 28 days following trial enrollment. Data are presented for the mITT population.
<sup>b</sup> These values were derived from Kaplan-Meier analysis.

#### Drug Acquisition and Administration Costs

A targeted literature review was conducted to support parameterization of costing inputs informing the model. All costs were reported in 2021 USD, with adjustments for inflation made based on the US Bureau of Labor Statistics Consumer Price Index for medical care, when appropriate [31]. The drug acquisition cost for lenzilumab was a hypothetical $10,000 per patient for the entire treatment course [32]. The treatment course for lenzilumab consisted of three 1-hour intravenous infusions, each administered 8 hours apart [13]. It was assumed that the cost to the hospital for each infusion was $34.87, for a total administration cost of $104.61. This cost was calculated based on the pharmacy labor and nonlabor (supplies) costs associated with administration of a monoclonal antibody over 60 minutes in a hospital-based setting, as reported by Schmier et al. [33]. Drug costs for SOC, including remdesivir and/or corticosteroids, were assumed to be captured in hospital resource use costs; they were not included as separate inputs in the model because it was assumed that lenzilumab would have no impact on the utilization or cost of background therapies. This assumption was supported by balanced remdesivir and corticosteroid use in both arms of the LIVE-AIR trial [13].

#### Hospital Resource Use and Costs

Inputs for hospital resource use included the proportion of patients and time to recovery for each level of care informed by data from the LIVE-AIR trial [19]. Inputs for daily hospital resource costs were obtained from the publication by Di Fusco et al. [8] by dividing the mean total hospital costs for each level of care by the corresponding mean length of stay. It was assumed that these daily costs incorporated all costs associated with SOC, including any remdesivir/corticosteroid use. Costs and resource use for each level of care are presented in **Table 2**.

**Table 2.**
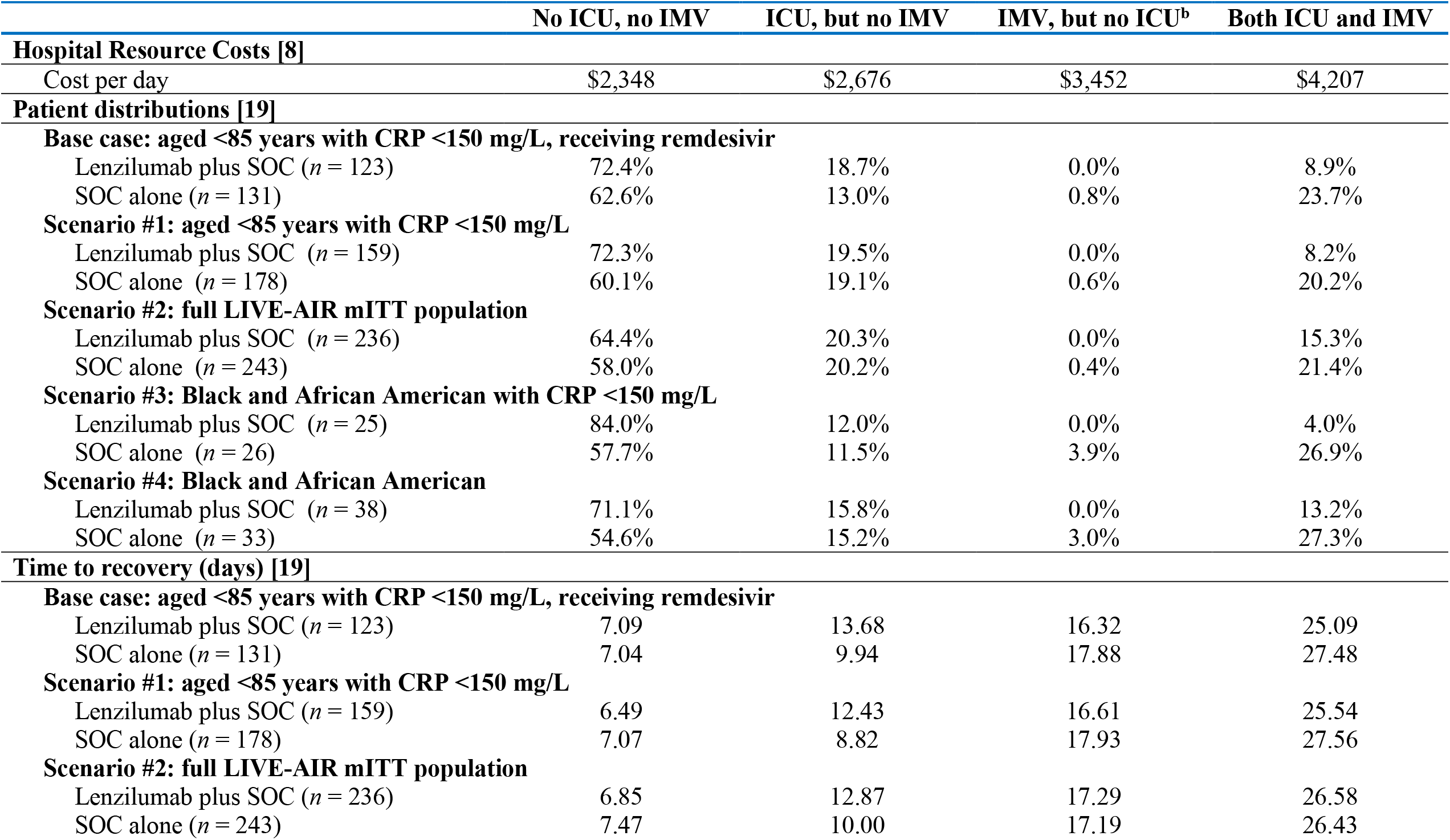
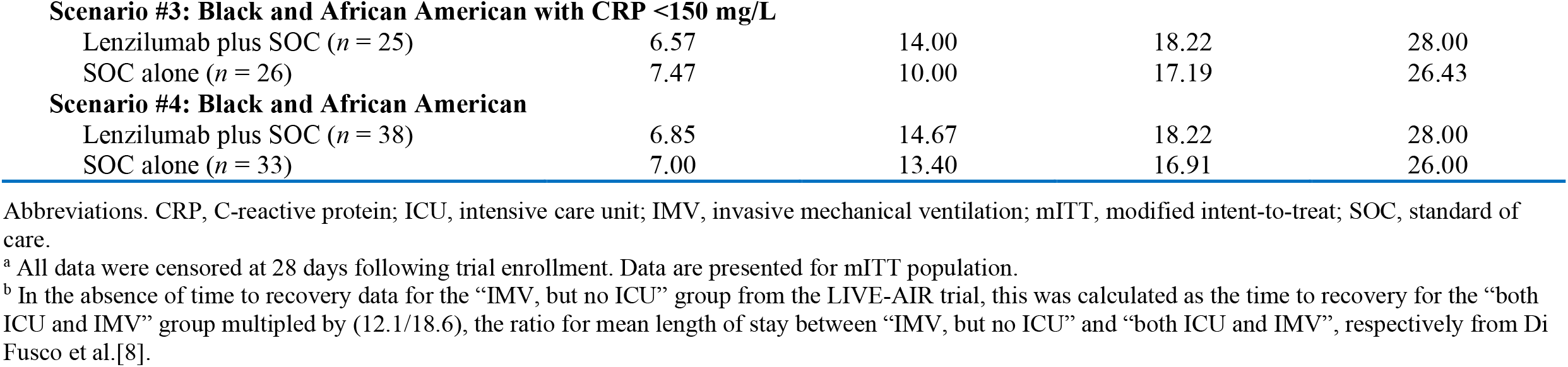
Hospital resource costs and use inputs from LIVE-AIR trial data^a^.

|  | No ICU, no IMV | ICU, but no IMV | IMV, but no ICU<sup>b</sup> | Both ICU and IMV |
| --- | --- | --- | --- | --- |
| **Hospital Resource Costs [8]** |  |  |  |  |
| Cost per day | \$2,348 | \$2,676 | \$3,452 | \$4,207 |
| **Patient distributions [19]** |  |  |  |  |
| **Base case: aged <85 years with CRP <150 mg/L, receiving remdesivir** |  |  |  |  |
| Lenzilumab plus SOC (*n* = 123) | 72.4% | 18.7% | 0.0% | 8.9% |
| SOC alone (*n* = 131) | 62.6% | 13.0% | 0.8% | 23.7% |
| **Scenario #1: aged <85 years with CRP <150 mg/L** |  |  |  |  |
| Lenzilumab plus SOC (*n* = 159) | 72.3% | 19.5% | 0.0% | 8.2% |
| SOC alone (*n* = 178) | 60.1% | 19.1% | 0.6% | 20.2% |
| **Scenario #2: full LIVE-AIR mITT population** |  |  |  |  |
| Lenzilumab plus SOC (*n* = 236) | 64.4% | 20.3% | 0.0% | 15.3% |
| SOC alone (*n* = 243) | 58.0% | 20.2% | 0.4% | 21.4% |
| **Scenario #3: Black and African American with CRP <150 mg/L** |  |  |  |  |
| Lenzilumab plus SOC (*n* = 25) | 84.0% | 12.0% | 0.0% | 4.0% |
| SOC alone (*n* = 26) | 57.7% | 11.5% | 3.9% | 26.9% |
| **Scenario #4: Black and African American** |  |  |  |  |
| Lenzilumab plus SOC (*n* = 38) | 71.1% | 15.8% | 0.0% | 13.2% |
| SOC alone (*n* = 33) | 54.6% | 15.2% | 3.0% | 27.3% |
| **Time to recovery (days) [19]** |  |  |  |  |
| **Base case: aged <85 years with CRP <150 mg/L, receiving remdesivir** |  |  |  |  |
| Lenzilumab plus SOC (*n* = 123) | 7.09 | 13.68 | 16.32 | 25.09 |
| SOC alone (*n* = 131) | 7.04 | 9.94 | 17.88 | 27.48 |
| **Scenario #1: aged <85 years with CRP <150 mg/L** |  |  |  |  |
| Lenzilumab plus SOC (*n* = 159) | 6.49 | 12.43 | 16.61 | 25.54 |
| SOC alone (*n* = 178) | 7.07 | 8.82 | 17.93 | 27.56 |
| **Scenario #2: full LIVE-AIR mITT population** |  |  |  |  |
| Lenzilumab plus SOC (*n* = 236) | 6.85 | 12.87 | 17.29 | 26.58 |
| SOC alone (*n* = 243) | 7.47 | 10.00 | 17.19 | 26.43 |
| **Scenario #3: Black and African American with CRP <150 mg/L** |  |  |  |  |
| Lenzilumab plus SOC (*n* = 25) | 6.57 | 14.00 | 18.22 | 28.00 |
| SOC alone (*n* = 26) | 7.47 | 10.00 | 17.19 | 26.43 |
| **Scenario #4: Black and African American** |  |  |  |  |
| Lenzilumab plus SOC (*n* = 38) | 6.85 | 14.67 | 18.22 | 28.00 |
| SOC alone (*n* = 33) | 7.00 | 13.40 | 16.91 | 26.00 |

Abbreviations. CRP, C-reactive protein; ICU, intensive care unit; IMV, invasive mechanical ventilation; mITT, modified intent-to-treat; SOC, standard of care.
<sup>a</sup> All data were censored at 28 days following trial enrollment. Data are presented for mITT population.
<sup>b</sup> In the absence of time to recovery data for the “IMV, but no ICU” group from the LIVE-AIR trial, this was calculated as the time to recovery for the “both ICU and IMV” group multiplied by (12.1/18.6), the ratio for mean length of stay between “IMV, but no ICU” and “both ICU and IMV”, respectively from Di Fusco et al.[8].

#### Rehospitalization Costs

Rehospitalizations within one year of the index COVID-19 hospitalization were included in the model as long-term costs and explored in scenario analysis #5 (in additional to the index hospitalization). Given the paucity of rehospitalization data specific to COVID-19 pneumonia, it was assumed that rates from a study of acute respiratory distress syndrome (ARDS) patients by Wu et al. [34] could be used as a proxy. The study included ARDS patients who received mechanical ventilation in their index hospitalization and non-ARDS hospitalized controls [34]. Thus, it was assumed that ARDS and non-ARDS patients from the study represented IMV and non-IMV patients in the model, respectively. Based on the study by Wu et al. [34], a rehospitalization rate of 53.2% was assumed for surviving IMV patients and 12.9% for non-IMV patients.

Rehospitalization costs were also derived from Wu et al. [34], with the assumption that not all of the annual long-term costs for healthcare resource use reported in the study were associated with rehospitalization. Based on a study of acute lung injury survivors by Ruhl et al. [35], it was assumed that 76% of the annual costs from Wu et al. [34] were from hospital readmissions. Therefore, it was assumed that if a patient was rehospitalized within one year of the index hospitalization, the cost would be $77,502 and $21,232 for IMV and non-IMV patients, respectively, after adjusting for inflation.

### Scenario Analyses

As mentioned previously, the base case analysis included patients aged <85 years with CRP <150 mg/L, and who were receiving remdesivir. Two scenario analyses were conducted to examine the budget impact of lenzilumab in broader patient populations. Scenario analysis #1 included patients aged <85 years with CRP <150 mg/L, with or without remdesivir. The full LIVE-AIR mITT population was included in scenario analysis #2. Two scenario analyses were also conducted to examine the budget impact in narrower patient populations. Scenario analysis #3 included Black and African American patients with CRP <150 mg/L, with or without remdesivir. Black and African American patients from the full mITT population were included in scenario analysis #4. Lastly, scenario analysis #5 added rehospitalization costs to the base case analysis to evaluate the combined budget impact to the hospital for the index hospitalization and estimated one-year long-term care costs.

### Sensitivity Analyses

Sensitivity analyses were conducted to determine the key budget impact drivers of the model. The following model inputs were included for sensitivity analyses: daily hospital costs, patient distributions in each level of care (lenzilumab plus SOC arm only), and time to recovery (lenzilumab plus SOC arm only). Inputs were adjusted by ±10%. Daily hospital costs for each level of care were adjusted simultaneously, as were time to recovery inputs. For patient distributions, the proportion of patients requiring both ICU and IMV was adjusted first and then the three other levels of care were reweighted, maintaining the proportionality between the levels of care prior to adjustment. The decision to use the ICU and IMV group for adjustment in the sensitivity analysis was based on the assumption that this group was the largest driver of costs in the model, as a result of the increased cost of care per day and the longer length of stay.

## Results

### Base Case and Scenario Analyses

In the base case and all scenario analyses, treatment with lenzilumab plus SOC improved all specified clinical outcomes over SOC alone (**Table 3**). In the base case, the number needed to treat (NNT) findings showed that every sixth treated patient avoided IMV or death if the cohort was treated with lenzilumab plus SOC rather than SOC alone. Across the scenario analyses this NNT ranged from a low of 4 in the Black and African American with CRP <150 mg/L subgroup up to 15 in the full LIVE-AIR mITT population. Lenzilumab also provided a benefit in terms of mortality, with an NNT of 10 to save one life in the base case (ranging from 8 to 23 across the scenario analyses). Ventilator use was also reduced by 15.5% (ranging from 6.5% to 26.8%) with the addition of lenzilumab to SOC compared with SOC alone and there were 3.90 IMV days saved (ranging from 1.85 up to 5.50 days). Finally, lenzilumab plus SOC improved the time to recovery, saving 2.40 bed days (range: 0.99 up to 4.92 days) as well as reducing time in ICU by 2.97 ICU days (range: 1.21 up to 5.06 days).

**Table 3.** Estimated clinical benefits of lenzilumab plus SOC over SOC alone per treated patient.

|  | Base case: aged <85<br>years with CRP <150<br>mg/L, receiving<br>remdesivir | Scenario #1: aged<br><85 years with CRP<br><150 mg/L | Scenario #2: full<br>LIVE-AIR mITT<br>population | Scenario #3: Black<br>and African<br>American with CRP<br><150 mg/L | Scenario #4: Black<br>and African<br>American |
| --- | --- | --- | --- | --- | --- |
| **Failure to achieve SWOV<sup>a</sup>** |  |  |  |  |  |
| Lenzilumab plus SOC | 9.2% | 8.4% | 15.6% | 4.0% | 13.2% |
| SOC alone | 24.8% | 21.2% | 22.1% | 29.3% | 29.1% |
| NNT for one patient to achieve SWOV | **6** | **8** | **15** | **4** | **6** |
| **Ventilator use** |  |  |  |  |  |
| Lenzilumab plus SOC | 8.9% | 8.2% | 15.3% | 4.0% | 13.2% |
| SOC alone | 24.4% | 20.8% | 21.8% | 30.8% | 30.3% |
| Reduced IMV use | **15.5%** | **12.6%** | **6.5%** | **26.8%** | **17.1%** |
| **Mortality<sup>a</sup>** |  |  |  |  |  |
| Lenzilumab plus SOC | 7.5% | 6.5% | 9.6% | 4.0% | 7.9% |
| SOC alone | 17.3% | 13.9% | 13.9% | 17.1% | 16.5% |
| NNT for one life saved | **10** | **14** | **23** | **8** | **12** |
| **Time to recovery (days)** |  |  |  |  |  |
| Lenzilumab plus SOC | 9.93 | 9.21 | 11.09 | 8.32 | 10.87 |
| SOC alone | 12.33 | 11.61 | 12.08 | 13.24 | 13.45 |
| Bed days saved | **2.40** | **2.40** | **0.99** | **4.92** | **2.58** |
| **Time in ICU (days)** |  |  |  |  |  |
| Lenzilumab plus SOC | 3.79 | 3.48 | 5.40 | 1.48 | 4.68 |
| SOC alone | 6.76 | 6.21 | 6.61 | 6.54 | 7.06 |
| ICU days saved | **2.97** | **2.73** | **1.21** | **5.06** | **2.38** |
| **Time on IMV (days)** |  |  |  |  |  |
| Lenzilumab plus SOC | 2.12 | 1.92 | 3.53 | 1.12 | 3.11 |
| SOC alone | 6.02 | 5.25 | 5.38 | 6.62 | 6.48 |
| IMV days saved | **3.90** | **3.33** | **1.85** | **5.50** | **3.37** |

Abbreviations. CRP, C-reactive protein; ICU, intensive care unit; IMV, invasive mechanical ventilation; mITT, modified intent-to-treat; NNT, number needed to treat; SOC, standard of care; SWOV, survival without ventilation.
<sup>a</sup> These values were derived from Kaplan-Meier analysis.

In addition to improved clinical outcomes, adding lenzilumab to SOC was also estimated to produce cost savings of $3,190 per patient in the base case analysis (**Table 4**). Scenario analysis #1 (aged <85 years with CRP <150 mg/L, with or without remdesivir) also estimated cost savings of $1,858 per patient. In scenario analysis #2 (full LIVE-AIR mITT population), addition of lenzilumab to SOC was estimated to result in a budget impact of $4,952 per patient over SOC alone. Scenario analysis #3 (Black and African American patients with CRP <150 mg/L, with or without remdesivir) and scenario analysis #4 (Black and African American patients from the full mITT population) estimated cost savings of $13,154 and $2,763 per patient, respectively. In addition to the index hospitalization, rehospitalization costs were included for scenario analysis #5 (with base case parameters for all other inputs); the addition of rehospitalization costs yielded further cost savings of $1,964 per patient in addition to the base case ($3,190), for total per patient cost savings of $5,154.

**Table 4.**
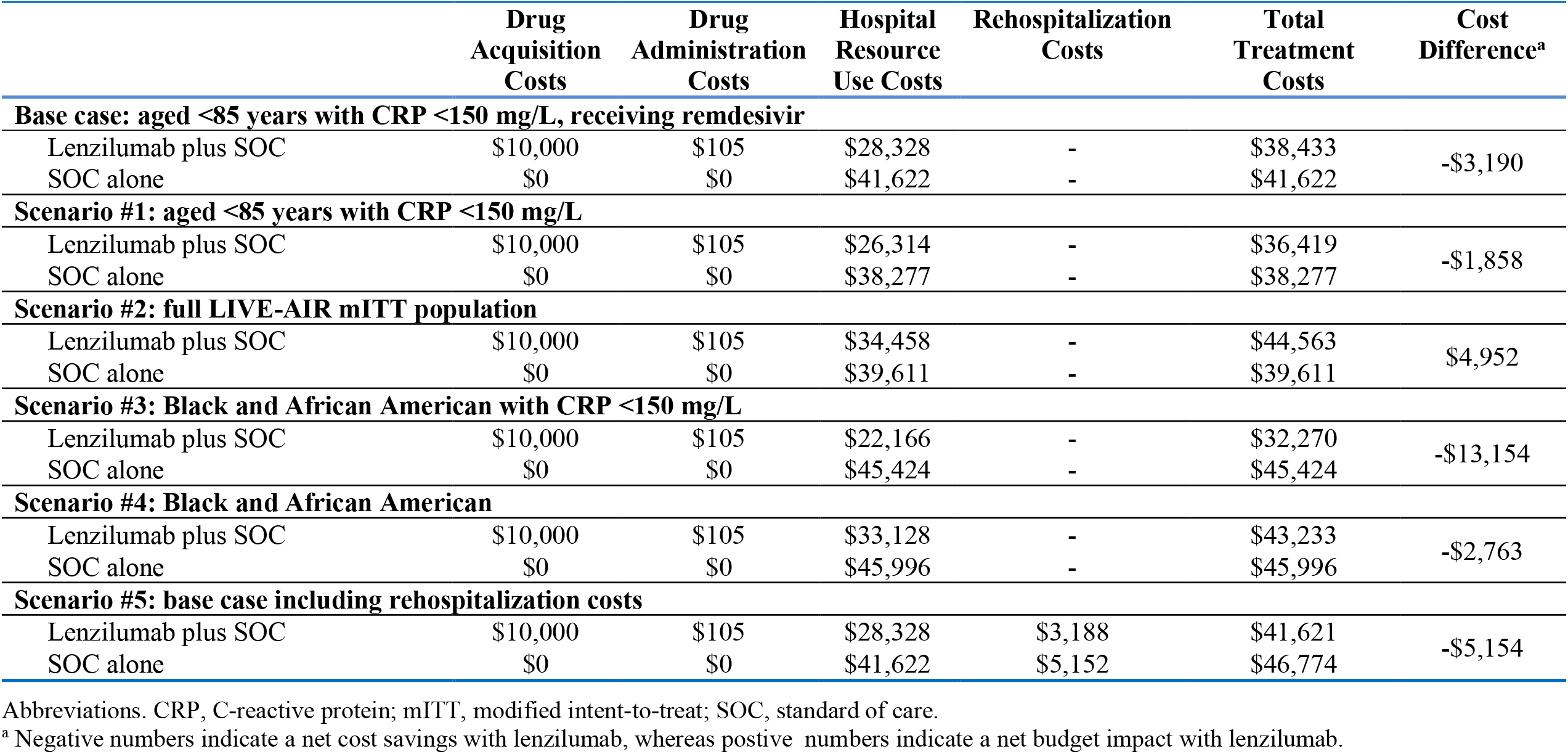
Estimated budget impact of lenzilumab plus SOC versus SOC alone per treated patient.

### Sensitivity Analyses

Inputs for hospital resource costs, patient distributions, and time to recovery were adjusted by ±10% (with base case parameters for all other inputs) in sensitivity analyses to determine the key budget impact model drivers. While all sensitivity analyses resulted in cost savings for lenzilumab plus SOC compared with SOC alone, there was variability relative to the base case results (**Table 5**).

**Table 5.**
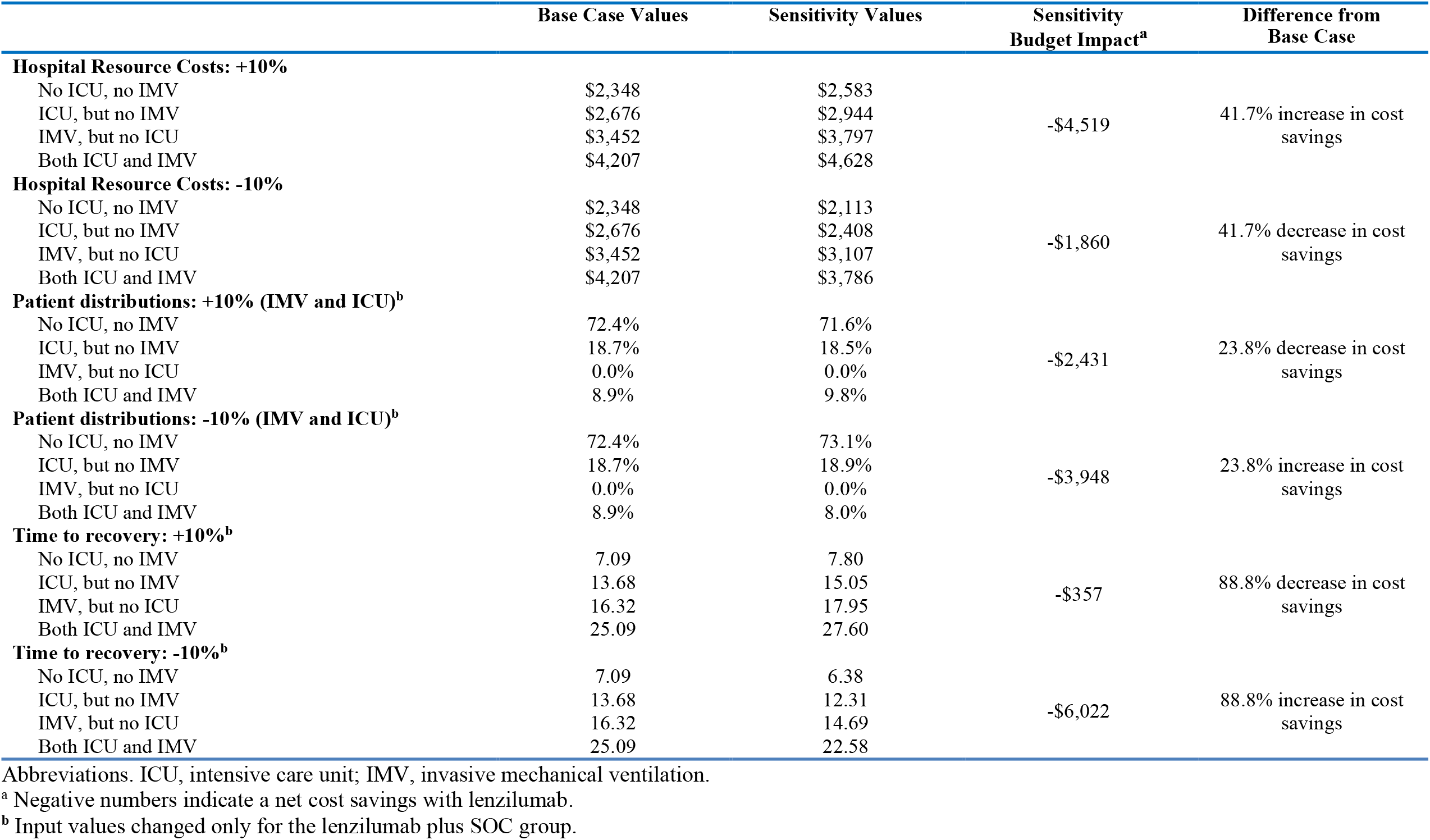
Sensitivity Analyses.

|  | Base Case Values | Sensitivity Values | Sensitivity Budget Impact<sup>a</sup> | Difference from Base Case |
| --- | --- | --- | --- | --- |
| **Hospital Resource Costs: +10%** |  |  |  |  |
| No ICU, no IMV | \$2,348 | \$2,583 | -\$4,519 | 41.7% increase in cost savings |
| ICU, but no IMV | \$2,676 | \$2,944 | | |
| IMV, but no ICU | \$3,452 | \$3,797 | | |
| Both ICU and IMV | \$4,207 | \$4,628 | | |
| **Hospital Resource Costs: -10%** |  |  |  |  |
| No ICU, no IMV | \$2,348 | \$2,113 | -\$1,860 | 41.7% decrease in cost savings |
| ICU, but no IMV | \$2,676 | \$2,408 | | |
| IMV, but no ICU | \$3,452 | \$3,107 | | |
| Both ICU and IMV | \$4,207 | \$3,786 | | |
| **Patient distributions: +10% (IMV and ICU)<sup>b</sup>** |  |  |  |  |
| No ICU, no IMV | 72.4% | 71.6% | -\$2,431 | 23.8% decrease in cost savings |
| ICU, but no IMV | 18.7% | 18.5% |  |  |
| IMV, but no ICU | 0.0% | 0.0% |  |  |
| Both ICU and IMV | 8.9% | 9.8% |  |  |
| **Patient distributions: -10% (IMV and ICU)<sup>b</sup>** |  |  |  |  |
| No ICU, no IMV | 72.4% | 73.1% | -\$3,948 | 23.8% increase in cost savings |
| ICU, but no IMV | 18.7% | 18.9% |  |  |
| IMV, but no ICU | 0.0% | 0.0% |  |  |
| Both ICU and IMV | 8.9% | 8.0% |  |  |
| **Time to recovery: +10%<sup>b</sup>** |  |  |  |  |
| No ICU, no IMV | 7.09 | 7.80 | -\$357 | 88.8% decrease in cost savings |
| ICU, but no IMV | 13.68 | 15.05 |  |  |
| IMV, but no ICU | 16.32 | 17.95 |  |  |
| Both ICU and IMV | 25.09 | 27.60 |  |  |
| **Time to recovery: -10%<sup>b</sup>** |  |  |  |  |
| No ICU, no IMV | 7.09 | 6.38 | -\$6,022 | 88.8% increase in cost savings |
| ICU, but no IMV | 13.68 | 12.31 |  |  |
| IMV, but no ICU | 16.32 | 14.69 |  |  |
| Both ICU and IMV | 25.09 | 22.58 |  |  |

Abbreviations. ICU, intensive care unit; IMV, invasive mechanical ventilation.
<sup>a</sup> Negative numbers indicate a net cost savings with lenzilumab.
<sup>b</sup> Input values changed only for the lenzilumab plus SOC group.

Varying the hospital resource costs resulted in cost savings ranging from $1,860 per patient with lower resource use costs to $4,519 per patient with higher resource use costs, a decrease and increase in cost savings of 41.7% relative to the base case, respectively. When decreasing the patient distribution in the IMV and ICU level of care for the lenzilumab plus SOC arm, the cost savings increased to $3,948 per patient, an increase in cost savings of 23.8% relative to the base. In contrast, increasing the patient distribution in the IMV and ICU decreased the cost savings by 23.8% relative to the base case to cost savings of $2,431 per patient. Finally, adjusting the time to recovery for the lenzilumab plus SOC arm ±10% had the most impact on the model results, with cost savings ranging from $357 per patient if time to recovery increased up to $6,022 per patient if time to recovery decreased, representing an 88.8% change in cost savings relative to the base case.

## Discussion

The present analysis evaluated the clinical benefits and budget impact of adding lenzilumab to SOC for the treatment of COVID-19 pneumonia from a US hospital perspective. The overall goal was to provide evidence that may assist hospital decision makers considering the use of lenzilumab as an option in the treatment of this condition that urgently requires new efficacious therapies.

Addition of lenzilumab to SOC was found to improve all specified clinical outcomes in the base case and scenario analyses, including SWOV, mortality, time to recovery, ICU use, and IMV use. SWOV is an important outcome among patients with COVID-19 from the hospital perspective, as avoiding IMV greatly reduces the average length of a patient’s hospital stay [36]. For all five analysis populations explored from the LIVE-AIR trial, the average time to recovery for patients with IMV was more than double that of patients without IMV [19]. In the present analysis, addition of lenzilumab to SOC resulted in a 15.5% absolute reduction in the probability of requiring IMV compared with SOC alone among patients aged <85 years with CRP <150 mg/L who were receiving remdesivir. The average number of bed days, ICU days, and IMV days were also reduced among patients who received lenzilumab in this patient population. Further, the NNT findings showed that every sixth patient avoided IMV or death if the cohort was treated with lenzilumab plus SOC rather than SOC alone. Lenzilumab also provided a benefit in terms of mortality, with an NNT of 10 to save one life. Therefore, for every 100 patients in this population, the results of this analysis suggest that 10 lives could be saved by treating with lenzilumab plus SOC in place of SOC alone.

Lenzilumab was also associated with a favorable budget impact from the hospital perspective in the base case analysis. Although the addition of lenzilumab to SOC was associated with increases in drug acquisition and administration costs compared with SOC alone, these additional costs were more than offset by the reduction in costs for hospital resource use among patients treated with lenzilumab. This led to a net per-patient cost savings of $3,190 with lenzilumab plus SOC compared with SOC alone among patients aged <85 years with CRP <150 mg/L who were receiving remdesivir.

In all scenario analyses conducted in both broader and narrower patient populations than that used for the base case analysis, addition of lenzilumab to SOC improved all clinical outcomes of interest compared with SOC alone. The relative clinical benefits were smaller in scenarios considering the broader populations (i.e., <85 years with CRP <150 mg/L with or without concomitant remdesivir and full mITT populations) than those observed in the base case population but greater in the subgroup analysis for Black and African American patients with CRP <150 mg/L. For the overall Black and African American population, the relative clinical benefit compared with the base case population varied across clinical outcomes.

In terms of budget impact, the addition of lenzilumab to SOC among patients aged <85 years with CRP <150 mg/L with or without concomitant remdesivir was still associated with a cost savings of $1,858, although this was smaller than the net savings in the base case population. When assessed in the full LIVE-AIR mITT population, the budget impact of adding lenzilumab to SOC was $4,952 per patient. Taken together, the results of these analyses show that adding lenzilumab to SOC for patients aged <85 years with CRP <150 mg/L, regardless of remdesivir use, may provide clinical and economic benefits for US hospitals. In a broader population of patients with COVID-19 pneumonia without consideration of age or CRP criteria, adding lenzilumab to SOC also improved clinical outcomes but with an increased budget impact.

Among patients hospitalized with COVID-19, elevated levels of CRP at the time of admission are positively correlated with disease severity and are associated with adverse clinical outcomes [13,20,21,37,38]. Further, CRP levels >100 mg/L at the time of admission are among the strongest independent predictors of critical illness [20,21], and patients with CRP levels >150 mg/L are considered at high risk for escalation of respiratory support (i.e., need for non-invasive ventilation or intubation) or death [39]. Given that lenzilumab resulted in particularly favorable clinical outcomes and cost savings among patients aged <85 years with CRP levels <150 mg/L, irrespective of whether all patients were receiving remdesivir, it appears to be most effective and provide the best economic value when used as an early intervention. Since testing for CRP levels is widely accessible and inexpensive in the US hospital setting [40], it may be a valuable and feasible approach to identify patients hospitalized with COVID-19 who may benefit most from treatment with lenzilumab.

Despite the Black and African American population being disproportionately affected by COVID-19, clinical trials frequently underrepresent this population [23]. As a result, there is a gap in knowledge regarding differences in disease severity, outcomes, and treatments across racial populations [27]. Results from a retrospective analysis of the LIVE-AIR trial suggest that Black and African American patients, in particular those with a CRP level <150 mg/L, demonstrate the greatest response to lenzilumab treatment [22]. In line with this, use of lenzilumab in Black and African American patients with CRP levels <150 mg/L with or without concomitant remdesivir resulted in cost savings per patient of $13,154, and a cost savings of $2,763 when all Black and African American patients from the LIVE-AIR mITT population were assessed. While findings from the Black and African American population in the LIVE-AIR trial are limited by the small sample size, it should be noted that the distribution of Black and African American patients within the clinical trial is proportional to the US ethnic distribution and thus, were well represented in the study overall [22,28].

Notably, the long-term health impacts of COVID-19 are not well characterized because of the novelty of the disease. However, the US CDC is monitoring several post-COVID conditions that may persist for weeks or months after initial infection and may impact most or all body systems [41]. Further, evidence from other recent outbreaks of conditions caused by similar coronaviruses, including severe acute respiratory syndrome (SARS) and Middle East respiratory syndrome (MERS), suggests that adverse respiratory, cardiovascular, and mental health outcomes may persist for months or years after resolution of the initial disease [42-46]. In terms of the present study, research into the long-term impact of COVID-19 would provide greater accuracy in the model’s budget impact estimations by reducing the number of assumptions required for rehospitalization costs. Despite these limitations, a scenario analysis was conducted to assess potential long-term cost savings with lenzilumab by incorporating rehospitalizations over one year after initial hospitalization into the analysis. The results indicated that adding lenzilumab to SOC may result in total cost savings of $5,154 associated with reduction in IMV use during the index hospitalization.

The sensitivity analyses suggested that model results were robust to changes reflecting a reasonable level of uncertainty (±10%) in key inputs including hospital resource costs, patient distributions to different levels of care, and time to recovery. Based on these analyses, time to recovery had the greatest impact on the results of the model. Decreasing the time to recovery by 10% for the lenzilumab plus SOC arm resulted in an 88.8% increase in cost savings relative to the base case. By comparison varying hospital resource costs and patient distributions to different levels of care had less impact on the cost savings associated with adding lenzilumab to SOC.

As with any model-based analysis, this study had several limitations. The analysis used data from the LIVE-AIR Phase 3 clinical trial, which would not be fully generalizable to a real-world US hospital population. Patient data from LIVE-AIR were censored after 28 days following trial enrollment. This is noteworthy because patients with COVID-19 who are critically ill and/or who require IMV typically have extended time to recovery, sometimes beyond 28 days [21,47,48]. Therefore, the model may underestimate time to recovery for patients who require IMV by only allowing up to 28 days of hospital resource use. Consequently, cost savings associated with the reduction in proportion of patients who require IMV may be even greater than suggested by the results of the current analysis. Finally, while the US Food and Drug Administration (FDA) recently (September 9^th^, 2021) declined the request for emergency use authorization (EUA) of lenzilumab to treat newly hospitalized COVID-19 patients based on the data from the LIVE-AIR trial concluding additional clinical data are required [49], the FDA has invited Humanigen to submit any supplemental data as they become available. It is also anticipated that the ongoing ACTIV-5/BET-B trial, which has been advanced to enroll up to 550 patients, may provide additional efficacy and safety data sufficient to further support the use of lenzilumab for the treatment of hospitalized COVID-19 patients [30].

Overall, the results of this analysis highlight the clinical benefits for SWOV, ventilator use, time to recovery, mortality, time in ICU, and time on IMV, in addition to a favorable budget impact from the US hospital perspective associated with adding lenzilumab to SOC for patients with COVID-19 pneumonia. Lenzilumab provides clinical benefits to a broad population of patients with characteristics similar to the full study cohort from the LIVE-AIR trial. Notably, the drug appears to be particularly effective in patients aged <85 years with CRP <150 mg/L and it is estimated to result in cost savings to US hospitals when used in this patient population. These clinical benefits and cost savings extend to the Black and African American subgroup as well. This is a critical finding as Black and African American persons are hyper-vulnerable to COVID-19 [23]. Concomitant use of remdesivir with lenzilumab further improves clinical benefits and cost savings. These findings support the use of lenzilumab as a standard option in the treatment of COVID-19 pneumonia and may help to assist hospital formulary decision makers with their consideration regarding its adoption in the hospital setting should an EUA or full approval through a Biologics License Application be granted.

## Data Availability

Some of the efficacy data used to inform the budget impact model was sourced from the Temesgen et al. (2021) publication (attached). Additional efficacy data are not publicly available at this time.

https://www.medrxiv.org/content/10.1101/2021.05.01.21256470v1

## Declaration of Funding

This work was supported by Humanigen Inc.

## Declaration of Financial/ Other Interests

AK, EJ, DC, MA are employees of Humanigen Inc. AZ, KT, ANP, AH, MT are employees of EVERSANA which received funding from Humanigen Inc. to conduct this study.

## Author Contributions

AK, EJ, AZ, KT, ANP, MT made substantial contributions to the conception and design of the model. Clinical data acquisition and analysis was conducted by DC. AZ, KT, ANP, AH conducted the literature review, developed the model structure, and prepared the manuscript. All authors reviewed the model and manuscript for interpretation of data and important intellectual content and have given their approval for the final version of the manuscript.

## Acknowledgements

The authors acknowledge Cameron Durant (Humanigen Inc.), Ari Mendell (EVERSANA), and Karson Theriault (EVERSANA) for their specific contributions to this project.

## Footnotes

1 SWOV is a robust composite endpoint used in many of the recent COVID-19 studies that is less prone to favor treatments with discordant effects on survival and days free of ventilation while avoiding the need for sample sizes approaching those of mortality trials to enable timely availability of study results.

2 Time to recovery was a pre-specified secondary outcome of the clinical trial and was defined as the first day on which a patient was discharged or ready for discharge by satisfying one of the following 3 categories from the 8-point ordinal scale: hospitalized, not requiring supplemental oxygen, no longer requiring ongoing medical care; not hospitalized, limitation on activities and/or requiring home oxygen; not hospitalized, no limitations on activities.

3 The mITT population was the analysis set used for the primary analysis of efficacy, defined as all randomized subjects who received at least one dose of study drug under the documented supervision of the principal investigator or sub-investigator and excluding sites that experienced documented limitations to access of basic supportive care for COVID-19.

## Notes

### Clinical Trial

NCT04351152

### Author Declarations

The current budget impact model was based on the The LIVE-AIR trial. Central IRB (ADVARRA) approval was obtained in addition to local IRB approval at each participating institution as appropriate for all aspects of this company-sponsored phase 3 clinical trial.

